# Correlation between adult tobacco smoking prevalence and mortality of Coronavirus Disease-19 across the world

**DOI:** 10.1101/2020.12.01.20241596

**Authors:** Nadya Magfira, Helda

## Abstract

**Background and aim:** Coronavirus disease 2019 (COVID-19) is a global pandemic spreading worldwide. Limited studies showed that smokers were at higher risk of having severe complications and higher mortality. This study aims to analyze the possible correlation between adult tobacco smoking prevalence and COVID-19 mortality all over the world.

**Methods:** This is a correlation study, we conducted a linear regression to analyse the correlation between smoking prevalence data in adults and COVID-19 Case Fatality Ratio (CFR) in countries with 1000 confirmed COVID-19 cases on May 3, 2020.

**Results:** Seventy-five country included with median CFR 3.66%. There are no relationships between adult male or female smoking prevalence with COVID-19 mortality in all over the countries. The multivariate analysis showed p-values of 0.823 and 0.910 for male and female smoking prevalence respectively. However, in lower-middle-income countries (LMIC), there is a positive correlation between the prevalence of adult male smoking with the lethality of COVID-19. Each percentage point increase in adult male smoking prevalence caused a CFR of COVID-19 increase by 0.08% (95% CI 0.00%-0.15%, p=0.041).

**Conclusions:** A correlation was found between the prevalence of adult male smoking and the CFR of COVID-19 in lower middle-income countries. Based on these findings, strengthening tobacco control policies is needed to reduce the impact of the COVID-19 pandemic especially in LMIC. Further researches are still needed.

## Introduction

Coronavirus Disease 2019 (COVID-19) is an infection caused by Severe Acute Respiratory Syndrome Coronavirus-2 (SARS CoV-2).^1^ COVID-19 has resulted in a public health emergency and an unprecedented economic crisis. On March 11, 2020, the World Health Organization (WHO) declared COVID-19 as a global pandemic. As of May 3^rd^, 2020 (the date of the latest data availability for this study) the disease has been infecting 3,349,786 people and causing 238,628 death.^2^

There is no clear light on when the COVID-19 pandemic will end. Studies show that it will take at least a decade to restore the social and economic conditions to their former state.^3^ Policies that effective, efficient, and provide broad effects on COVID-19 impact need to consider, one of it was applying social restriction. Following those policy, the time spent indoors are lengthened and those made the risk of harmful use of tobacco smoking increased.^4^

As we all know, smoking is main common risk factor of morbidity and mortality of various non-communicable diseases. We have also known that COVID-19 fatalities are higher among people with pre-existing conditions.^5^ In a study that predicted the 90-day mortality rate from viral pneumonias in hospitalized patients, smokers were twice as likely to die compared with those who did not smoke.^6^ Recent studies show smoking increases the risk of developing severe symptoms and increases the risk of in-hospital mortality. A meta-analysis by Zhao et al. showed that there was a significant relationship between smoking and the severity of COVID-19, OR 2.0 (95% CI 1.3-3.1).^7^ Studies by Zheng et.al and Guo et.al. also showed the same, smoking increased the risk of severe symptoms and mortality by twofold compared with those who did not smoke.^8 9^

By contrast, some researchers suggest a protective effect of smoking and COVID-19 based on epidemiologic data that were not controlled for age and comorbidities.^10^ Preliminary meta-analysis based on Chinese patients suggest that active smoking does not apparently seem to be significantly associated with enhanced risk of progressing towards severe disease in COVID-19.^11^ However, all studies included in this meta-analysis were limited in China.

In response to recent published possibility of smokers paradox, the centers for disease control and prevention and the World Health Organization recommend against smoking to reduce the risk of harm from the disease.^12^ Knowing that COVID-19 is a newly identified disease, the link between tobacco smoking and COVID-19 needs further research. Here we describe the correlation of adult tobacco smoking prevalence and the mortality of COVID-19 in all over the world.

## Methods

This is a correlation study using pooled data from all over the world. To investigate the correlation between adult smoking prevalence and COVID-19 mortality, only countries who had 1000 total confirmed cases by May 3rd, 2020 included. A minimum confirmed case was required because extremely low case may reflect inadequate testing. Countries with no available data on adult smoking prevalence were excluded from this study.

## 1. Data source

### Mortality Data

Data of COVID-19 cases and death per country were obtained from the Coronavirus disease (COVID-2019) situation report-104 by WHO (May 3rd, 2020) available from http://www.who.int/docs/default-source/coronaviruse/situation-reports/20200503-covid-19-sitrep-104.pdf?sfvrsn=53328f46_2.

### Variables

We collected data prevalence of tobacco smoking in male and female adults across countries from WHO situation data in 2020 available from https://apps.who.int/gho/data/node.main.65. The data of current health expenditure (% of Gross Domestic Product) obtained from The World Bank data available from https://data.worldbank.org/indicator/SH.XPD.CHEX.GD.ZS. The Prevalence of population ages 65 and above also obtained from The Worl Bank data available from https://data.worldbank.org/indicator/SP.POP.65UP.TO.ZS. The Prevalence of Diabetes was based on International Diabetes Atlas year 2019.^13^ The prevalence of hypertension and obesity were obtained from the latest WHO Global Health Observatory and Repository available from https://apps.who.int/gho/data/node.main.A867?lang=en. Countries’ income was classified according to world bank data available at https://datahelpdesk.worldbank.org/knowledgebase/articles/906519-world-bank-country-and-lending-groups. All data accessed on May 3^rd^, 2020.

### 2. Analysis

Case Fatality Ratio for each country was measured by dividing total deaths to the total confirmed case in percent. Attack rate was measured by dividing total confirmed case/ 100,000 population. We performed spearman correlation analysis to analyze the correlation between adult tobacco smoking prevalence and mortality of COVID-19. Multivariate analysis was conducted by performing linear regression analysis to evaluate the relationship incorporating the studied variables (attack rate, health expenditure, prevalence of population above 65, and comorbidity). Many factors might influence the CFR of COVID-19 including a country’s standard of medical care.^14^ To account for that we performed sub-analysis according to country Gross National Income per capita in 2018. We divided countries into two categories: low and lower-middle-income countries (LMIC) with an income below 3.995 dollars and upper-middle (UMIC) to high-income countries (HIC) with an income above 3.995 dollars.

## Results

By May 3rd, 2020, 89 countries had more than 1000 total confirmed cases of COVID-19. However, only 75 countries whose data on adult tobacco smoking prevalence available. This study includes 15 (20%) LMIC, 22 (29.33%) UMIC, and 38 (50.67%) HIC (Appendix 1). By that, a total of 3.229.365 confirmed cases of COVID-19 with 235,524 death cases enrolled. Our study shows that the median of CFR per country in May 3^rd^ 2020 was 3.66% (IQR: 3.89, min: 0.10 max: 19.10). The attack rate per 100,000 people was 65.78 (IQR: 153.28, min: 1.19, max: 614.94) and the median of adult male and female smoking prevalence was 30.3% (IQR: 20.2, min:8 max:82.7) and 12.6% (IQR: 17.7, min: 0.2 max: 39.3) respectively (table 1).

**Table 1.** Characteristic of Country.

|  | LMIC<br>N=15 (20%) | UMIC and HIC<br>N=60 (80%) | Total N=75 (100%) |
| --- | --- | --- | --- |
| Case Fatality Ratio (%) – median<br>(IQR), (n=75) | 3.08 (3.38) | 3.97 (4.08) | 3.66 (3.89) |
| Attack Rate/ 100,000 – median (IQR),<br>(n=75) | 7.13 (5.28) | 93.55 (167.97) | 65.78 (153.28) |
| Adult Tobacco Smoking Prevalence |  |  |  |
| Male – median (IQR), (n=75) | 39 (27.3) | 29.05 (19.8) | 30.3 (20.2) |
| Female – median (IQR), (n=75) | 1.2 (5.0) | 15.5 (15.5) | 12.6 (17.7) |
| Health expenditure (%) – median<br>(IQR), (n=75) | 4.45 (3.15) | 8.10 (3.49) | 7.10 (3.89) |
| Prevalence of population ≥ 65 (%) –<br>median (IQR), (n=75) | 5.28 (4.2) | 16.17 (8.57) | 14.67 (11.78) |
| Comorbidity |  |  |  |
| Prevalence of diabetes (%) –<br>median (IQR), (n=74) | 6.85 (2.7) | 8.35 (4.85) | 8.05 (4.2) |
| Prevalence of obesity (%) –<br>median (IQR), (n=73) | 10.5 (13.2) | 22.5 (5.8) | 22 (6.2) |
| Prevalence of hypertension (%) –<br>median (IQR), (n=75) | 22 (5.2) | 25.0 (10.7) | 23.5 (9.2) |

*LMIC: Low Middle-Income Countries, UMIC: Upper middle-income countries, HIC: high income countries, IQR: interquartile range*

From all countries included our study show significant negative correlation between the prevalence of adult male smoking and CFR of COVID-19. The Spearman’s rank correlation coefficient was −0.2645 (p=0.022, Figure 1A). We also found there is significant positive correlation between the prevalence of adult female smoking and CFR of COVID-19. The Spearman’s rank correlation coefficient was 0.3005 (p=0.009, Figure 1B).

**Figure 1.**
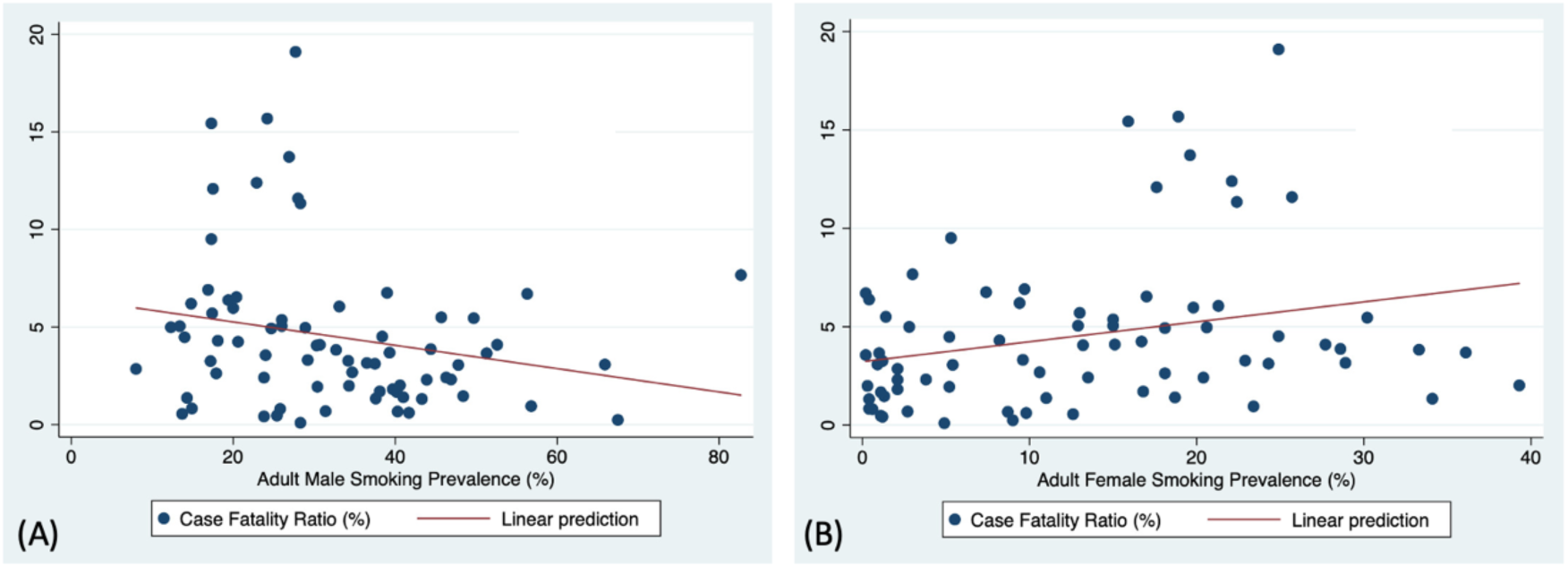
(A). Correlation of adult male smoking prevalence with COVID-19 case fatality ratio from all across countries (Spearman correlation r= −0.2645, p=0.022). (B). Correlation of adult female smoking prevalence with COVID-19 case fatality ratio from all across countries (Spearman correlation r=0.3005 p=0.009).

Table 2 and Table 3 shows linear regression analysis evaluating the relationship between adult male and adult female smoking prevalence with CFR of COVID-19 incorporating the studied variables. There are no significant relationships seen between adult male (p=0.823, table 2) and female smoking prevalence (p=0.910, table 3) with CFR of COVID-19 when including the confounding variable.

**Table 2.** Linear Regression Analyses of Male Smoking Prevalence and CFR of COVID-19.

|  | Total |  | LMIC |  | UMIC/HIC |  |
| --- | --- | --- | --- | --- | --- | --- |
| | $\beta \pm SE$ | P-value | $\beta \pm SE$ | P-value | $\beta \pm SE$ | P-value |
| Male Smoking | **$-0.01 \pm 0.03$** | **0.82** | **$0.07 \pm 0.03$** | **0.04\*** | **$-0.06 \pm 0.05$** | **0.16** |
| Prevalence (%) |  |  |  |  |  |  |
| Health Expenditure (%) | $0.23 \pm 0.25$ | 0.35 | $0.27 \pm 0.37$ | 0.50 | $0.25 \pm 0.27$ | 0.37 |
| Prevalence of | $0.17 \pm 0.13$ | 0.20 | $0.30 \pm 0.21$ | 0.21 | $0.21 \pm 0.16$ | 0.21 |
| population $\geq 65$ (%) | | | | | | |
| Prevalence of diabetes | $-0.28 \pm 0.14$ | 0.06 | $0.46 \pm 0.19$ | 0.05 | $-0.31 \pm 0.17$ | 0.07 |
| (%) |  |  |  |  |  |  |
| Prevalence of obesity | $0.03 \pm 0.06$ | 0.69 | $-0.10 \pm 0.02$ | 0.25 | $0.06 \pm 0.09$ | 0.50 |
| (%) |  |  |  |  |  |  |
| Prevalence of | $-0.06 \pm 0.10$ | 0.58 | $-0.44 \pm 0.18$ | 0.05 | $-0.02 \pm 0.12$ | 0.89 |
| hypertension (%) |  |  |  |  |  |  |
| Attack Rate/ 100,000 | $0.01 \pm 0.00$ | 0.21 | $0.2 \pm 0.40$ | 0.40 | $0.01 \pm 0.00$ | 0.25 |

LMIC: Low Middle-Income Countries, UMIC: Upper middle-income countries, HIC: high income countries, SE: standard error, \*) significant for $p$ -value $< 0.05$

Table 2. Linear Regression Analyses of Female Smoking Prevalence and CFR of COVID-19
|  | Total |  | LMIC |  | UMIC/HIC |  |
| --- | --- | --- | --- | --- | --- | --- |
| | $\beta \pm SE$ | P-value | $\beta \pm SE$ | P-value | $\beta \pm SE$ | P-value |
| Female Smoking Prevalence (%) | **0.01 ± 0.07** | **0.91** | **0.09 ± 0.32** | **0.80** | **-0.02 ± 0.08** | **0.85** |
| Health Expenditure (%) | 9.24 ± 0.25 | 0.33 | 0.02 ± 0.54 | 0.98 | 0.29 ± 0.27 | 0.30 |
| Prevalence of population ≥ 65 (%) | 0.17 ± 0.14 | 0.24 | 0.11 ± 0.68 | 0.87 | 0.23 ± 0.19 | 0.20 |
| Prevalence of diabetes (%) | -0.29 ± 0.14 | 0.05* | 0.50 ± 0.28 | 0.13 | -0.36 ± 0.17 | 0.03* |
| Prevalence of obesity (%) | 0.03 ± 0.07 | 0.71 | -0.05 ± 0.11 | 0.69 | 0.08 ± 0.09 | 0.37 |
| Prevalence of hypertension (%) | -0.07 ± 0.11 | 0.54 | -0.30 ± 0.42 | 0.50 | -0.04 ± 0.13 | 0.73 |
| Attack Rate/ 100,000 | 0.01 ± 0.00 | 0.19 | 0.02 ± 0.03 | 0.53 | 0.01 ± 0.00 | 0.12 |

LMIC: Low Middle-Income Countries, UMIC: Upper middle-income countries, HIC: high income countries, SE: standard error, \*) significant for p-value < 0.05

**Figure 2.**
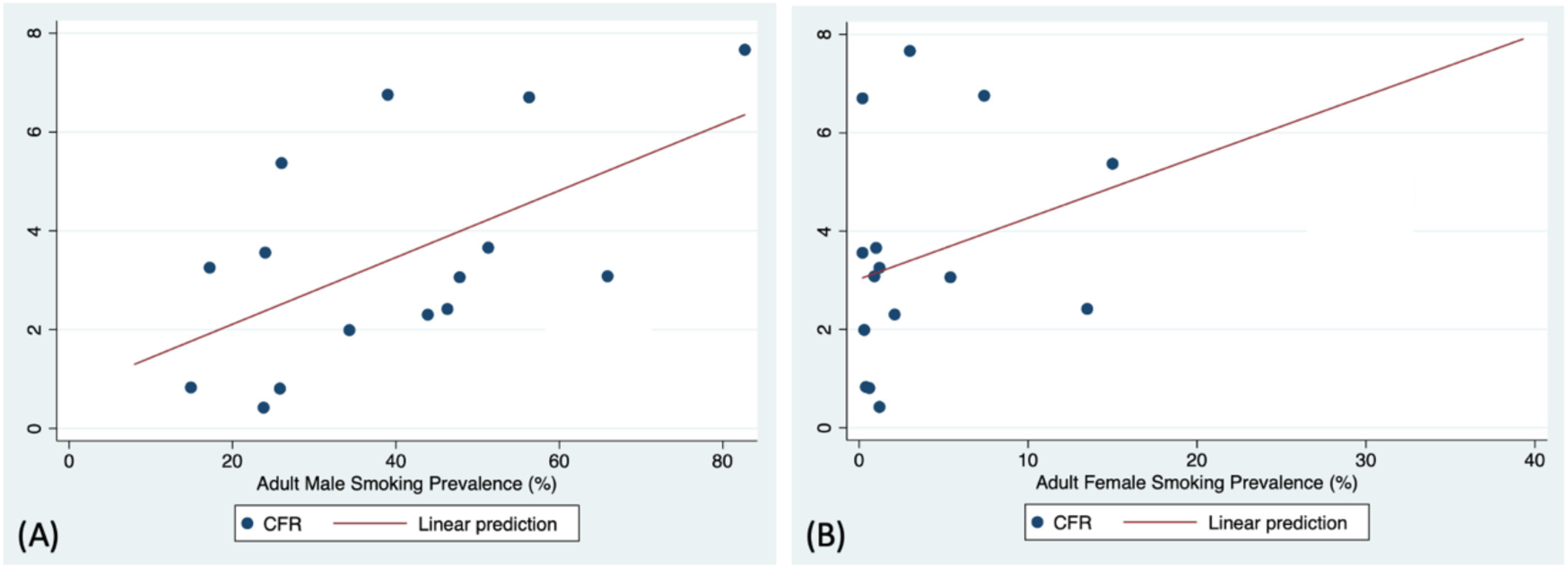
(A) Correlation of adult male smoking prevalence with COVID-19 case fatality ratio in Lower middle-income group (Spearman correlation r=0.5214, p=0.046). (B). Correlation of adult female smoking prevalence with COVID-19 case fatality ratio in Lower middle-income group (Spearman correlation r=0.2272, p=0.415).

**Figure 3.**
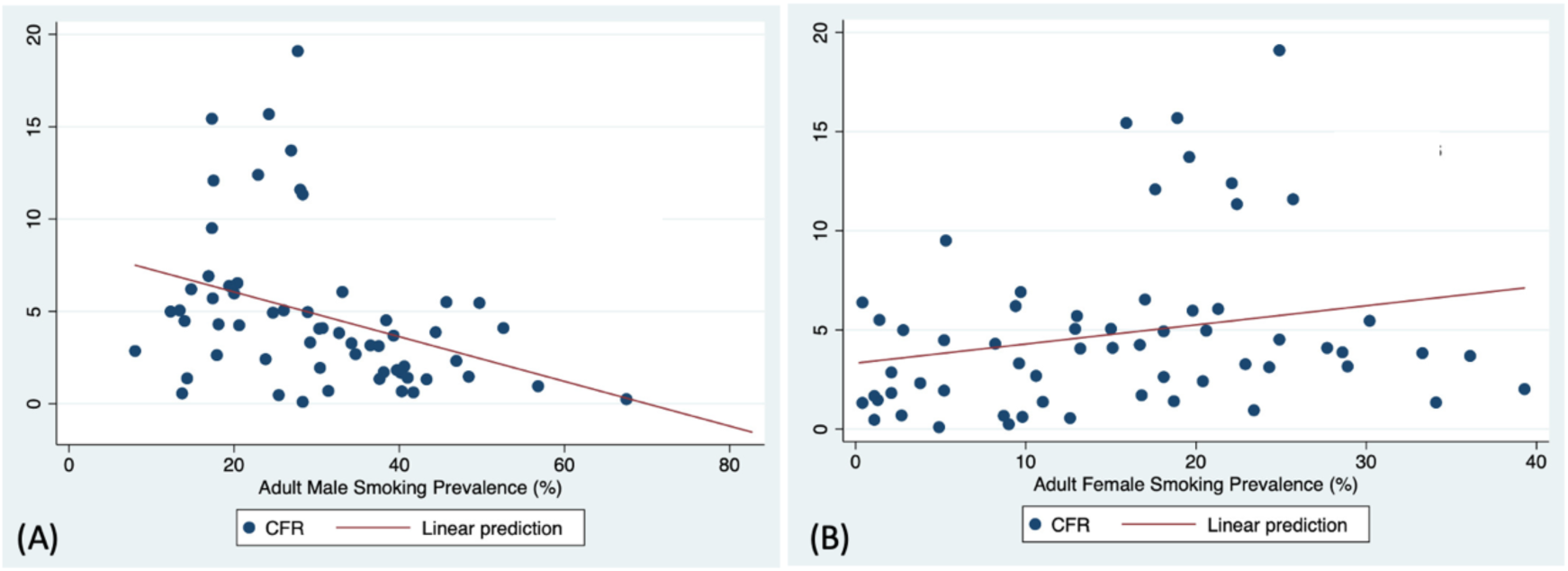
(A) Correlation of adult male smoking prevalence with COVID-19 case fatality ratio in upper and high-income countries group (Spearman correlation r=-0.4472, p=0.000). (B). Correlation of adult female smoking prevalence with COVID-19 case fatality ratio in upper and high-income countries group (Spearman correlation r=0.2706, p=0.037).

### a. Correlation Between Adult Tobacco Smoking and CFR of COVID-19 in LMIC

In LMIC, we found a significant positive correlation between the prevalence of adult male smoking and CFR of COVID-19. The Spearman’s rank correlation coefficient was 0.5214 (p=0.046). Each percentage point increase in adult male smoking prevalence caused a COVID-19 CFR increase of 0.08% (95% CI 0.00%-0.15%, p=0.041) in LMIC (table 2). However, we don’t find any correlation between the prevalence of adult female smoking and CFR of COVID-19. The Spearman’s rank correlation coefficient was 0.2272 (p=0.415).

### b. Correlation Between Adult Smoking and CFR of COVID-19 in UMIC and HIC

In UMIC and HIC, we found a significant negative correlation between the prevalence of adult male smoking and CFR of COVID-19. The Spearman’s rank correlation coefficient was −0.4472 (p=0.000). We also found there is significant positive correlation between the prevalence of adult female smoking prevalence and CFR of COVID-19. The Spearman’s rank correlation coefficient was 0.2706 (p=0.037). However, there are no significant relationship seen between adult male and female smoking prevalence with CFR of COVID-19 in UMIC and HIC when including the confounding variable (table 2).

## Discussion

To best of our knowledge, this is the most comprehensive study to analyse the correlation of adult female and male smoking prevalence with the lethality of COVID-19 in all over the world using pooled data from 75 countries. The global spread of tobacco use has been long known to have an impact on human health. However, only few studies available explaining how tobacco impacts COVID-19 patients.

Our study showed there are no relationships between adult male or female smoking prevalence with COVID-19 mortality in all over the countries. The multivariate analysis showed p-values of 0.823 and 0.910 for male and female smoking prevalence respectively. However, in lower-middle-income countries (LMIC), there is a positive correlation between the prevalence of adult male smoking with the lethality of COVID-19. Each percentage point increase in adult male smoking prevalence caused a CFR of COVID-19 increase by 0.08% (95% CI 0.00%-0.15%, p=0.041).

Our study has some potential limitation to be discussed, first, we received the data of COVID-19 from situation reports by WHO, the number of deaths and cases might not always reflect the exact situation since there are differences in how different Governments across countries identifying infected cases. Second, in our report, we performed sub-analysis by categorizing countries according to their GNI per capita. However, there are still many factors that can influence the lethality of COVID-19 in which we do not control. Thus, including prevalence of other related comorbidities such as chronic kidney disease, heart disease, stroke, respiratory disease, malignancy and autoimmune conditions. We also did not include weather parameters situation of each countries which may contribute to COVID-19 CFR.^15^

In bivariate analysis, we found a significant negative correlation between prevalence of adult male smoking and CFR of COVID-19 in all over the world also in sub-analysis UMIC and HIC groups. However, after adjusting with other variables there are no significant correlations found. Some studies reported active smokers are under represented among COVID-19 patients and there is also a study that show protective effect of smoking in COVID-19 mortality.^16 17^ Those studies lead to widespread claim that smoking maybe protective against COVID-19.^17^ However, knowing that early in pandemic there was a race to publish, this is likely resulted in aberrant and non-standardized data collection and poor statistical analysis which can lead to erroneous conclusion. Both of the studies were not having quite variables to control the relationship. Reports of the protective effects of smoking on COVID-19 are unfounded. There are no anyway the results of previous studies be an indicator to start or continue smoking.

In 2015, it was estimated that 80% of smokers all over the world live in the low-and low-middle-income-countries.^18^ Our study showed that in LMIC there is a positive correlation between the prevalence of adult male smoking with COVID-19 mortality. The result was also significant after adjustment with other variables. Each percentage point increase in adult male smoking prevalence caused CFR of COVID-19 increase by 0.08% (95% CI 0.00%-0.15%, p=0.041).

Some mechanisms on how smoking interact with COVID-19 outcomes are; smokers have increased gene expression of Angiotensin Enzyme 2 (ACE2), a known receptors of SARS CoV-2 than previous smokers and non-smokers.^19^ It leads to an increase in vasoconstriction, vascular permeability, inflammation, and acute lung injury.^20^ There is also evidence more circulating ACE2 in men which provides evidence for gender-based variations in disease severity.^21^ Smoking increase the risk of lung damage by destroying ciliated epithelium and disrupts its function which protects the lungs through the production of mucus and rapid clearance of pathogens.^19 22^ Smoking has been shown to up-regulate inflammation through activation of nuclear factor kappa-light-chain-enhancer of activated B cells, tumor necrosis factor-a, IL-1beta, and neutrophils. Smoking has been reported to down-regulate CXCL-10, a chemokine that is important for the recruitment of macrophages, neutrophils and natural killer cells, minimizing the capacity of the innate immune system to suppress viral replication.^23^

In LMIC, due to the rising population including a large youth population, growing incomes and prosperity, and relatively poor tobacco control, the prevalence of adult male smoking was higher compared to UMIC/HIC.^18^ Other than that, the results might be influenced by several factors like limited testing of COVID-19 in the LMIC group. This can be proven from mean attack rate value which is only 7.13 at LMIC while at UMIC/HIC it can reach 93.55. Also, there are different capacity of the country to managed the disease that could influenced the outcome. In HIC the policies on tobacco control is well established.^18^ The battle against tobacco use should continue, no other risk factors are as immediately modifiable as smoking. Gas exchange, lung function, and blood circulation, improve quickly after smoking cessation.^19^ Strengthening tobacco control policies and assisting smokers to successfully and permanently quit are needed.

## Conclusion

In conclusion, a correlation was found between the prevalence of adult male smoking and the CFR of COVID-19 in LMIC. Based on these findings, strengthening tobacco control policies and assisting smokers to successfully and permanently quit are needed to reduce the impact of the COVID-19 pandemic especially in LMIC.

## Data Availability

The data that support the findings of this study are openly available in WHO situation data, WHO Global Health Observatory and Repository, and The World Bank data

https://apps.who.int/gho/data/node.main.65.

https://data.worldbank.org/indicator/SH.XPD.CHEX.GD.ZS.

https://apps.who.int/gho/data/node.main.A867?lang=en.

## APPENDIX 1

| Country | GNI | Confirmed Case | Death | CFR (%) | Male smoking (%) | Female smoking (%) |
| --- | --- | --- | --- | --- | --- | --- |
| Bangladesh | LMIC | 8790 | 175 | 1,99 | 34,3 | 0,3 |
| Bolivia (Plurinational State of) | LMIC | 1229 | 66 | 5,37 | 26 | 15 |
| Cameroon | LMIC | 2077 | 64 | 3,08 | 65,9 | 0,9 |
| Egypt | LMIC | 6193 | 415 | 6,70 | 56,3 | 0,2 |
| Ghana | LMIC | 2169 | 18 | 0,83 | 14,9 | 0,4 |
| India | LMIC | 39980 | 1301 | 3,25 | 17,2 | 1,2 |
| Indonesia | LMIC | 10843 | 831 | 7,66 | 82,7 | 3 |
| Morocco | LMIC | 4729 | 173 | 3,66 | 51,3 | 1 |
| Nigeria | LMIC | 2388 | 85 | 3,56 | 24 | 0,2 |
| Pakistan | LMIC | 19103 | 440 | 2,30 | 43,9 | 2,1 |
| Philippines | LMIC | 8928 | 603 | 6,75 | 39 | 7,4 |
| Republic of Moldova | LMIC | 4052 | 124 | 3,06 | 47,8 | 5,4 |
| Senegal | LMIC | 1115 | 9 | 0,81 | 25,8 | 0,6 |
| Ukraine | LMIC | 11913 | 288 | 2,42 | 46,3 | 13,5 |
| Uzbekistan | LMIC | 2127 | 9 | 0,42 | 23,8 | 1,2 |
| Argentina | UMIC | 4532 | 229 | 5,05 | 26 | 15 |
| Armenia | UMIC | 2386 | 35 | 1,47 | 48,4 | 1,3 |
| Azerbaijan | UMIC | 1894 | 25 | 1,32 | 43,3 | 0,4 |
| Belarus | UMIC | 15828 | 97 | 0,61 | 41,7 | 9,8 |
| Bosnia and Herzegovina | UMIC | 1757 | 68 | 3,87 | 44,4 | 28,6 |
| Brazil | UMIC | 91589 | 6329 | 6,91 | 16,9 | 9,7 |
| Bulgaria | UMIC | 1594 | 72 | 4,52 | 38,4 | 24,9 |
| China | UMIC | 84393 | 4643 | 5,50 | 45,7 | 1,4 |
| Colombia | UMIC | 7006 | 314 | 4,48 | 14 | 5,2 |
| Cuba | UMIC | 1611 | 66 | 4,10 | 52,6 | 15,1 |
| Dominican Republic | UMIC | 7578 | 326 | 4,30 | 18,1 | 8,2 |
| Ecuador | UMIC | 27464 | 1371 | 4,99 | 12,3 | 2,8 |
| Iran (Islamic Republic of) | UMIC | 96448 | 6156 | 6,38 | 19,4 | 0,4 |
| Kazakhstan | UMIC | 3877 | 26 | 0,67 | 40,3 | 8,7 |
| Malaysia | UMIC | 6176 | 103 | 1,67 | 40,2 | 1,1 |
| Mexico | UMIC | 20739 | 1972 | 9,51 | 17,3 | 5,3 |
| Romania | UMIC | 12732 | 771 | 6,06 | 33,1 | 21,3 |
| Russian Federation | UMIC | 134687 | 1280 | 0,95 | 56,8 | 23,4 |
| Serbia | UMIC | 9362 | 189 | 2,02 | 40,6 | 39,3 |
| South Africa | UMIC | 6336 | 123 | 1,94 | 30,4 | 5,2 |
| Thailand | UMIC | 2969 | 54 | 1,82 | 39,7 | 2,1 |
| Turkey | UMIC | 124375 | 3336 | 2,68 | 34,7 | 10,6 |
| Australia | HIC | 6783 | 93 | 1,37 | 14,3 | 11 |
| Austria | HIC | 15558 | 596 | 3,83 | 32,7 | 33,3 |
| Bahrain | HIC | 3284 | 8 | 0,24 | 67,5 | 9 |
| Belgium | HIC | 49517 | 7765 | 15,68 | 24,2 | 18,9 |
| Canada | HIC | 55572 | 3446 | 6,20 | 14,8 | 9,4 |
| Chile | HIC | 18435 | 247 | 1,34 | 37,6 | 34,1 |
| Croatia | HIC | 2088 | 77 | 3,69 | 39,3 | 36,1 |
| Czechia | HIC | 7755 | 245 | 3,16 | 36,5 | 28,9 |
| Denmark | HIC | 9407 | 475 | 5,05 | 13,4 | 12,9 |
| Estonia | HIC | 1699 | 53 | 3,12 | 37,5 | 24,3 |
| Finland | HIC | 5179 | 220 | 4,25 | 20,6 | 16,7 |
| France | HIC | 129458 | 24724 | 19,10 | 27,7 | 24,9 |
| Germany | HIC | 162496 | 6649 | 4,09 | 30,7 | 27,7 |
| Greece | HIC | 2620 | 143 | 5,46 | 49,7 | 30,2 |
| Hungary | HIC | 2998 | 340 | 11,34 | 28,3 | 22,4 |
| Iceland | HIC | 1798 | 10 | 0,56 | 13,7 | 12,6 |
| Ireland | HIC | 21176 | 1265 | 5,97 | 20 | 19,8 |
| Israel | HIC | 16152 | 227 | 1,41 | 41 | 18,7 |
| Italy | HIC | 209328 | 28710 | 13,72 | 26,9 | 19,6 |
| Japan | HIC | 14839 | 492 | 3,32 | 29,2 | 9,6 |
| Lithuania | HIC | 1406 | 46 | 3,27 | 34,2 | 22,9 |
| Luxembourg | HIC | 3812 | 92 | 2,41 | 23,8 | 20,4 |
| Netherlands | HIC | 40236 | 4987 | 12,39 | 22,9 | 22,1 |
| Norway | HIC | 7759 | 204 | 2,63 | 17,9 | 18,1 |
| Oman | HIC | 2568 | 12 | 0,47 | 25,4 | 1,1 |
| Panama | HIC | 6720 | 192 | 2,86 | 8 | 2,1 |
| Poland | HIC | 13375 | 664 | 4,96 | 28,9 | 20,6 |
| Portugal | HIC | 25190 | 1023 | 4,06 | 30,3 | 13,2 |
| Republic of Korea | HIC | 10793 | 250 | 2,32 | 46,9 | 3,8 |
| Saudi Arabia | HIC | 25459 | 176 | 0,69 | 31,4 | 2,7 |
| Singapore | HIC | 17548 | 17 | 0,10 | 28,3 | 4,9 |
| Slovakia | HIC | 1407 | 24 | 1,71 | 38,1 | 16,8 |
| Slovenia | HIC | 1439 | 94 | 6,53 | 20,4 | 17 |
| Spain | HIC | 216582 | 25100 | 11,59 | 28 | 25,7 |
| Sweden | HIC | 22082 | 2669 | 12,09 | 17,5 | 17,6 |
| Switzerland | HIC | 29734 | 1466 | 4,93 | 24,7 | 18,1 |
| The United Kingdom | HIC | 182264 | 28131 | 15,43 | 17,3 | 15,9 |
| United States of America | HIC | 1093880 | 62406 | 5,71 | 17,4 | 13 |

## Notes

### Competing Interest Statement

The authors have declared no competing interest.

### Funding Statement

This research did not receive any specific grant from funding agencies in the public, commercial, or not for profit sector

### Author Declarations

The study used public use data set, I confirm all relevant ethical guidelines have been followed and no necessary IRB approval needed

## References

1. Lu H, Stratton CW, Tang YW. Outbreak of pneumonia of unknown etiology in Wuhan, China: The mystery and the miracle. J Med Virol 2020;92(4):401–02. doi: 10.1002/jmv.25678 [published Online First: 2020/01/18]

2. Organization WH. Coronavirus disease (COVID-19) Situation Report – 104. World Health Organization 2020

3. Djalante R, Lassa J, Setiamarga D, et al. Review and analysis of current responses to COVID-19 in Indonesia: Period of January to March 2020. Progress in Disaster Science 2020;6 doi: 10.1016/j.pdisas.2020.100091

4. Kluge HHP, Wickramasinghe K, Rippin HL, et al. Prevention and control of non-communicable diseases in the COVID-19 response. The Lancet 2020;395(10238):1678–80. doi: 10.1016/s0140-6736(20)31067-9

5. Wang B, Li R, Lu Z, et al. Does comorbidity increase the risk of patients with COVID-19: evidence from meta-analysis. Aging (Albany NY) 2020;12(7):6049–57. doi: 10.18632/aging.103000 [published Online First: 2020/04/09]

6. Guo L, Wei D, Zhang X, et al. Clinical Features Predicting Mortality Risk in Patients With Viral Pneumonia: The MuLBSTA Score. Frontiers in Microbiology 2019;10 doi: 10.3389/fmicb.2019.02752

7. Zhao Q, Meng M, Kumar R, et al. The impact of COPD and smoking history on the severity of COVID-19: A systemic review and meta-analysis. J Med Virol 2020 doi: 10.1002/jmv.25889 [published Online First: 2020/04/16]

8. Zheng Z, Peng F, Xu B, et al. Risk factors of critical & mortal COVID-19 cases: A systematic literature review and meta-analysis. J Infect 2020;81(2):e16–e25. doi: 10.1016/j.jinf.2020.04.021 [published Online First: 2020/04/27]

9. Guo FR. Smoking links to the severity of COVID-19: An update of a meta-analysis. J Med Virol 2020 doi: 10.1002/jmv.25967 [published Online First: 2020/05/06]

10. Changeux JP, Amoura Z, Rey FA, et al. A nicotinic hypothesis for Covid-19 with preventive and therapeutic implications. C R Biol 2020;343(1):33–39. doi: 10.5802/crbiol.8 [published Online First: 2020/07/29]

11. Lippi G, Henry BM. Active smoking is not associated with severity of coronavirus disease 2019 (COVID-19). Eur J Intern Med 2020;75:107–08. doi: 10.1016/j.ejim.2020.03.014 [published Online First: 2020/03/21]

12. Marquizo AB. Tobacco control during the COVID-19 pandemic: how we can help. WHO Farmework convention on tobacco control 2020

13. Federation ID. IDF Diabetes Atlas Ninth Edition 2019. International Diabetes Federation 2019

14. Miller A, Reandelar MJ, Fasciglione K, et al. Correlation between universal BCG vaccination policy and reduced mortality for COVID-19. 2020 doi: https://doi.org/10.1101/2020.03.24.20042937

15. Ma Y, Zhao Y, Liu J, et al. Effects of temperature variation and humidity on the death of COVID-19 in Wuhan, China. Science of The Total Environment 2020;724 doi: 10.1016/j.scitotenv.2020.138226

16. Williamson EJ, Walker AJ, Bhaskaran K, et al. Factors associated with COVID-19-related death using OpenSAFELY. Nature 2020;584(7821):430–36. doi: 10.1038/s41586-020-2521-4 [published Online First: 2020/07/09]

17. Usman MS, Siddiqi TJ, Khan MS, et al. Is there a smoker’s paradox in COVID-19? BMJ Evid Based Med 2020 doi: 10.1136/bmjebm-2020-111492 [published Online First: 2020/08/14]

18. Anderson C, Becher H, Winkler V. Tobacco Control Progress in Low and Middle Income Countries in Comparison to High Income Countries. International Journal of Environmental Research and Public Health 2016;13(10) doi: 10.3390/ijerph13101039

19. Silva ALOd, Moreira JC, Martins SR. COVID-19 e tabagismo: uma relação de risco. Cadernos de Saúde Pública 2020;36(5) doi: 10.1590/0102-311X00072020

20. Lang AE, Yakhkind A. Coronavirus Disease 2019 and Smoking: How and Why We Implemented a Tobacco Treatment Campaign. Chest 2020 doi: 10.1016/j.chest.2020.06.013 [published Online First: 2020/06/21]

21. Kaur G, Lungarella G, Rahman I. SARS-CoV-2 COVID-19 susceptibility and lung inflammatory storm by smoking and vaping. J Inflamm (Lond) 2020;17:21. doi: 10.1186/s12950-020-00250-8 [published Online First: 2020/06/13]

22. Simet SM, Sisson JH, Pavlik JA, et al. Long-Term Cigarette Smoke Exposure in a Mouse Model of Ciliated Epithelial Cell Function. American Journal of Respiratory Cell and Molecular Biology 2010;43(6):635–40. doi: 10.1165/rcmb.2009-0297OC

23. Todt JC, Freeman CM, Brown JP, et al. Smoking decreases the response of human lung macrophages to double-stranded RNA by reducing TLR3 expression. Respir Res 2013;14:33. doi: 10.1186/1465-9921-14-33 [published Online First: 2013/03/19]

